# Reflective Writing as a Medical Education Intervention in Value Identification

**DOI:** 10.1101/2025.08.14.25333710

**Authors:** Shelby L. McCubbin, Raven R. Piercey

## Abstract

Values are an essential aspect of day-to-day life, both in healthcare and in society as a whole. Values affect both the smaller decisions we make and our overall goals. Because of this, it is important for physicians to be prepared to help patients identify individual values, which begins with needing to be able to identify their own personal values. One way to prepare students to do this is through the lens of narrative medicine, a discipline which uses the skills of close reading and reflective writing as techniques for enhancing clinical skills and patient care. An introductory workshop on reflective writing and value identification was offered to first- and second-year medical students. The workshop was capped at twelve participants to ensure a small group environment. Participants were provided with a brief presentation on value identification, followed by a guided reflection time. Students were presented with a passage from a physician memoir, followed by three reflective writing prompts. After the reflective writing portion, participants shared their writing and discussed the value of community while in healthcare training and how reflective writing groups help reduce burnout and create better connection between peers. Workshop members also highlighted the ways in which these skills could apply to their future practice in a variety of different fields (including primary care, surgical, and further specialized). In fact, 92% of participants marked “Strongly Agree” for the statements: “This workshop was relevant to my future as a physician.” and “An understanding of personal values will help me to be a better future physician.” The combined metrics of the optional end of workshop survey as well as the remarks made during group discussion lead to the conclusion that small reflective writing groups focused around a specific topic offer a valuable communication and relationship intervention in medical education.

## Introduction

Narrative medicine is one of the most studied fields within the medical humanities (Palla 2024). Coined by Rita Charon, a scholar at Columbia University, narrative medicine relies on “narrative skills of recognizing, absorbing, interpreting, and being moved by the stories of illness” (Charon 2001). The expanse of narrative medicine is not easy to summarize. It consists of many different practices (such as close reading, reflective writing) and a wide multitude of academic disciplines (art, literature, etc.). This paper hones in on the benefits that reflective writing practices can provide to early career medical students, particularly in the context of value identification and specification of future goals.

Reflective writing serves as an active process in which students individually assess a familiar or recent event, including an acknowledgement of not only what happened, but also any emotions or biases related to said event (Zannini 2018). This practice can present in various forms within medicine, with one common method being parallel charting. Parallel charting is designed to supplement typical chart notes that medical professionals are familiar with. Parallel charts are written in plain language (instead of medical jargon) and aim to describe not only the encounter itself, but also how the encounter impacted the provider/student. Research indicates that students who engage in parallel charting gain a better understanding of their patients and form enhanced patient-provider relationships (Banfi et al. 2018). Besides offering physicians and students a chance to reflect on their experiences and lessons learned from specific encounters, reflective writing also enables students to share their perspectives and experiences with others.

Medical students have a unique perspective on healthcare, functioning as both outsiders and insiders within the field. As students transition from pre-clinicals to clinicals, often during the third year of medical school, they are far enough into their training that they have learned what questions to ask when collecting a history and appropriate physical exam maneuvers. Many students have begun to perfect their “script” with patients, and may even start to feel comfortable walking into a patient’s room. Patients see this, and label medical students as “insiders.” However, to residents, attendings, and anyone else higher up in the chain of command, medical students may still be seen as outsiders. Though students have finished their preclinical curriculum and are officially training to practice, they do not yet fully understand the terminology, expectations, or rituals associated with the profession. This level of understanding will only come with hands-on experience as students continue to progress in their training. With continued training and experience, we see an eventual transition from medical student as perceived outsider to acceptance as true insider. This imbalance and eventual transition in roles is addressed through professional socialization, which involves internalizing the profession’s culture, identifying its values, and incorporating these values into their behavior (Zarshenas et al., 2014). Professional socialization is essential for integrating students into the larger professional community and fostering a sense of belonging (Forourzadeh et al. 2018).

Various techniques are believed to aid in professional socialization, with storytelling being a particularly effective method (Erogul 2022) This then allows us to see a connection between the importance of professional socialization and the practice of reflective writing, as both storytelling and reflective writing involve reflecting on events to identify learned lessons. Thus, reflective writing can be seen as a form of storytelling, suggesting that it may facilitate professional socialization. Engaging in reflective writing can help medical students feel more at ease during their transition into practice after medical school, and the intimacy of sharing writing can foster better relationships, not only among peers, but also between students and their supervisors (Shapiro 2006).

An essential aspect of professional socialization is the integration of a profession’s values into a new professional’s personal life. Previous research has found a bidirectional relationship between a new trainee’s acquisition of values (and other aspects of a workplace’s culture) and professional socialization outcomes (Miller 2010). It is also recognized that values are a large part of our everyday life. Values are considered to be the “transsituational goals serving as guiding principles in a person’s life” (Nilsson and Lundmark 2020). Personal values affect the learning styles a student adopts (Gamage et al 2021), the prioritization of competing goals (Schwartz 2010), and the standards upon which we evaluate people and events (Schwartz 1992). Values are a part of most of our everyday lives, and yet we spend little time focusing on them or refining our own personal values. In fact, there is a great lack in the literature of how to teach students to identify their own personal values, or even the benefit of doing so. This workshop therefore aims to help bridge this perceived gap.

## Materials and Methods

### Workshop design and model

The workshop began with a 30-minute introduction to narrative medicine and reflective writing methods. Because the workshop was advertised to medical students, regardless of experience with the medical humanities, it was essential to create a standard knowledge base for everyone to work off. The information presented in this component of the workshop was summarized from personal thesis work done several years prior centered around medical memoirs as an educational resource (McCubbin 2022). Students were first given background information on what the medical humanities are, and more specifically foundational principles of narrative medicine. They were then given an overview of communication in medicine as well as the concept of narrative competence. This led to a brief overview on the practice of reflective writing, as well as an introduction to the memoir *When Breath Becomes Air* by Paul Kalanithi.

Paul Kalanithi completed his residency at Stanford in Neurosurgery. However, he also had a background in literature, as well as in the history and philosophy of science and medicine. In his sixth year of residency, Kalanithi was diagnosed with stage 4 non-small-cell EGFR-positive lung cancer. His book focused on learning of his diagnosis and having to figure out how to navigate his own mortality. This memoir is unique from many other medical memoirs as it is an example of the same person sharing both a patient and physician perspective. It was deemed necessary for inclusion in the workshop because of a particular passage where Kalanithi describes his own values and how they have changed.

> “The tricky part of illness is that, as you go through it, your values are constantly changing. You try to figure out what matters to you, and then you keep figuring it out. It felt like someone had taken away my credit card and I was having to learn how to budget. You may decide you want to spend your time working as a neurosurgeon, but two months later, you may feel differently. Two months after that, you may want to learn to play the saxophone or devote yourself to the church. Death may be a one-time event, but living with terminal illness is a process.” (Kalanithi 160-161).

This passage shows medical students the relevance of values to their practice and lives as future physicians, and was thus able to transition the workshop from a place of theory to a time of practice.

At this point, students were guided through a set of three prompts, each of which was designed to highlight a students’ personal core values to them in abstract ways. These prompts were pulled from a theoretical educational activity designed around the above quote (McCubbin 2022). The three prompts were:

- Who do you admire?
- When have you taken action?
- When do you feel most at home (authentic)?

For each prompt, the question was read aloud and then students were given three minutes to write and respond. They were given a warning when approximately 10-15 seconds were remaining so that they knew to finish up their thoughts. At the conclusion of each prompt, participants were given the chance to read their work to the group. This was caveated by saying that no justification, explanation, or excuses were allowed to accompany the reading. The facilitator first read their response aloud each time, and then the floor was open to anyone else who wanted to participate. No one was required to share at any point during the workshop, though most participants chose to read at least once.

After the completion of the reflective component of the workshop, the students were given a brief presentation on personal values and how they affect our day to day lives. This then led into a 15-20 minute group discussion addressing the following questions:

- Why is it important to reflect on our own values?
- Did responding to these three prompts at all change the way you think about what matters most to you?
- How could completing this exercise better prepare students to help future patients refine their own core values?

### Participants

The workshop was advertised to first- and second-year medical students at the University of Kentucky College of Medicine. This cohort of students were chosen as they are at the earliest level of career advancement (and thus professional socialization), yet they are still committed to a career in medicine as indicated by acceptance to medical school. Participation was restricted to the first twelve students to RSVP to attend. We intentionally wanted a small group of students to create an intimate environment where they were more likely to feel comfortable sharing their responses to prompts. Participants included ten first-year medical students and two second-year medical students. As this was a voluntary workshop and not meant to represent the entire school, we did not collect any further demographic information on participants.

### Workshop assessment

At the end of the workshop, students were given the chance to fill out a survey rating how much they agreed or disagreed with a set of statements. Each participant chose to fill out the survey.

The statements included were:

- I can apply skills from this workshop when working with patients.
- The collaborative environment of the workshop contributed to a beneficial experience.
- The workshop was relevant to my future as a physician.
- Reflective writing is an important tool for physicians in **any** field.
- An understanding of personal values will help me to be a better future physician.
- I think other students would benefit from participating in this workshop.
- I would participate in a similar workshop again.

Each statement was accompanied by a 5 point Likert scale where students could mark an answer ranging from “Strongly agree” to “Strongly disagree”. After rating agreement/disagreement with the seven statements, there was an area where participants could comment on strengths or weaknesses of the workshop.

### Data analysis

To assess reliability and confirm that the various items were intercorrelated enough to be combined into a scale, an alpha coefficient was calculated using the software SPSS and found to be 0.701. This indicates relatively high internal consistency and that the items can be grouped to measure student perception of workshop success and relevance. As this was an optional education intervention, we did not collect extensive demographic information or divide participants into groups. Because of this, all tests were run as one-sample. Though it was proven that the data could be grouped into a single scale, we still ran a one-sample binomial test for each item (besides item 7 for which a one-sample chi-square test was required) to determine whether there was a significant difference between survey responses. With this being an optional workshop, we knew that responses were more likely to be skewed in favor of the workshop. But by completing the one-sample binomial tests, we were able to determine whether there was an equal likelihood of participants selecting “Strongly Agree” versus “Agree”. With the final comments section (strengths and weaknesses of the workshop) being optional, several participants chose not to say anything. That said, the comments that were left were coded for common themes.

## Results

Twelve students participated in a 1.5-hour pilot reflective writing workshop. Students were not required to have any humanities background or reflective writing experience coming in. Of the twelve students, ten were first-year medical students and two were second-year medical students. Everyone stayed for the full session, and all twelve participants completed the optional end of workshop survey.

The stacked bar graph allows a collective visualization of responses to the various survey measures. Each blue bar represents students who strongly agreed with the statement, red marks agreed, and yellow marks neutral. We did not have any participant mark disagree or strongly disagree for any of the items. It can therefore be concluded that overall assessment of the workshop was favorable. As noted in the methods, an alpha coefficient of 0.701 allows us to treat these individual items as part of one larger scale indicating attitudes toward the workshop.

**Table 1.**
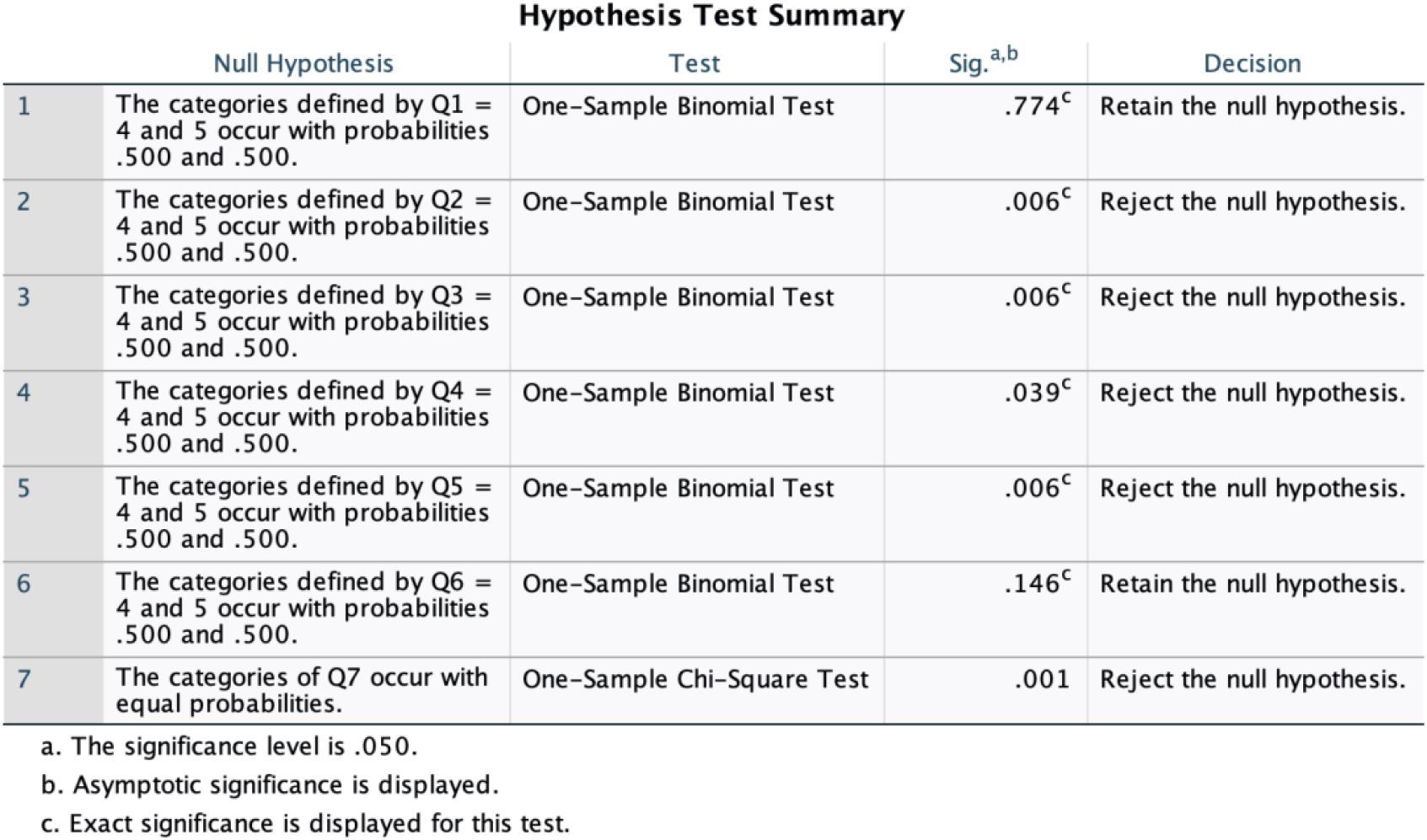
Hypothesis Test Summary. Each line of the table indicates the difference between the number of people who chose a particular response for that particular survey item. Statistical analysis showed a significant number of people selecting “Strongly agree” for items 2, 3, 4, 5, and 7 (as listed in order in Figure 1).

**Figure 1.**
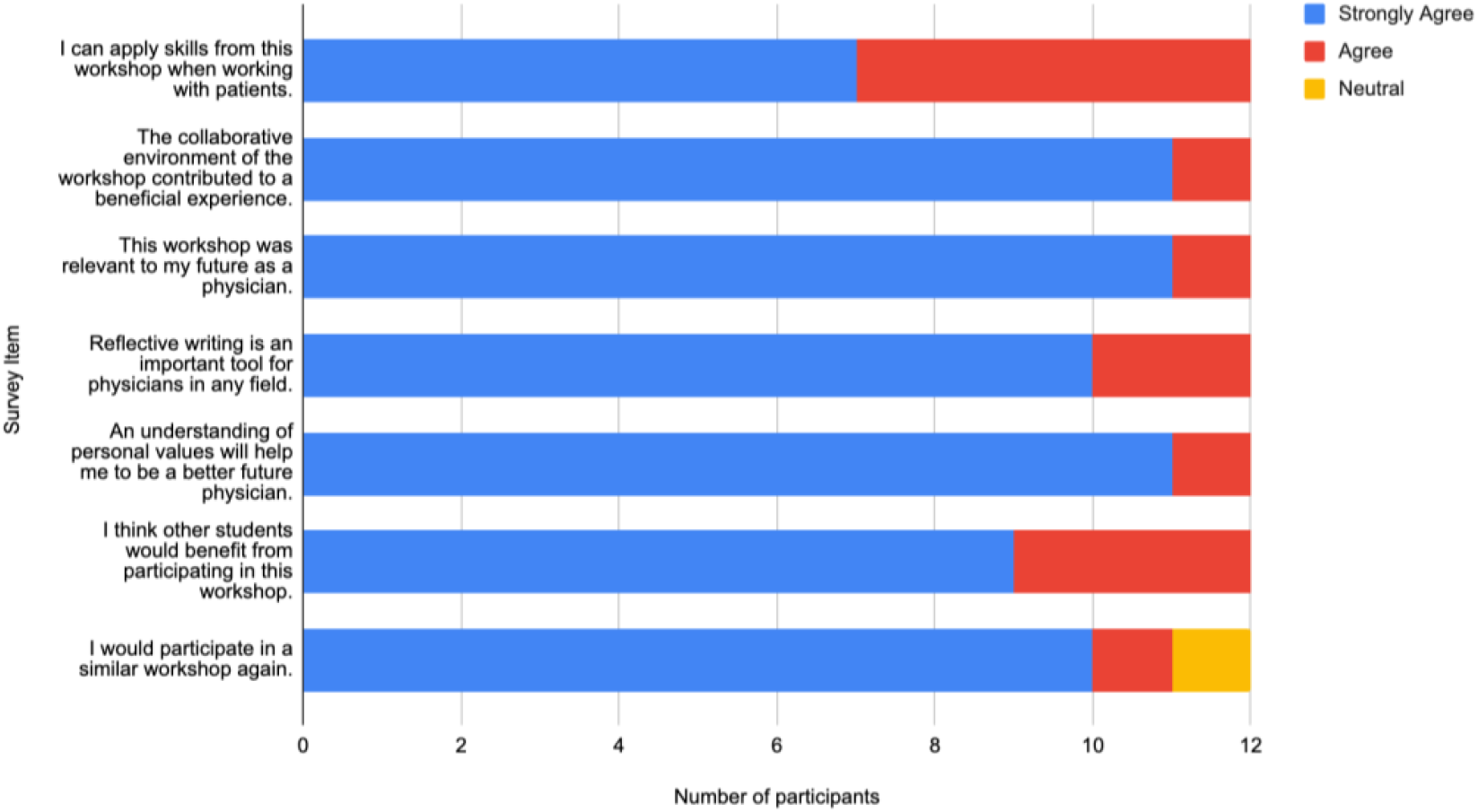
Perceived Benefit of Workshop. Each survey item is listed as a separate bar so that overall perception of the workshop can be visualized. If each individual survey item is grouped into a larger Likert scale of “Perceived benefit” (which can be done due to calculated Cronbach’s alpha), it could be concluded that the majority strongly agree.

Though overall assessment was favorable, we wanted to determine whether there was a significant difference between participants’ tendency to mark “Agree” versus “Strongly agree”. As the workshop was voluntary, it was not surprising for most students to mark “Agree”. However, a higher likelihood of “Strongly agree” could suggest higher benefit in participation for future groups. One-Sample binomial tests indicate that there is in fact a significant difference for the items: “The collaborative environment of the workshop contributed to a beneficial experience.”, “This workshop was relevant to my future as a physician.”, “Reflective writing is an important tool for physicians in any field.”, “An understanding of personal values will help me to be a better future physician.”, and “I would participate in a similar workshop again.” (p=0.006, 0.006, 0.039, 0.006, 0.001 respectively).

When looking at optional strength/weakness comments, 33% of the total participants commented specifically on the benefits of the collaborative/conversational nature of the workshop. 42% of participants noted that this process is important and a nice break from traditional biomedical curriculum. Other individual highlights were the benefits of vulnerability, creation of a space for gratitude, and the benefit of connecting with like minded individuals. The only weakness commented on was the reflection time. 17% of participants noted in their written comments that they wish there was more time to write to the prompts, and several other participants brought this up during the informal discussion at the end of the workshop.

## Discussion

This optional workshop offered preclinical students the opportunity to learn about reflective writing and value identification. Students did not know the specific topic of the workshop ahead of time, leading to a more authentic reaction to chosen prompts. At the conclusion of the session, participants were asked to complete an optional end-survey. All participants chose to complete it, and overall expressed strong perceived benefit of the workshop.

One of the greatest benefits of this intervention was that students were highly engaged throughout the workshop and were able to develop community with the other participants. We did not ask participants ahead of time whether they knew each other, but being from two different medical classes and a variety of different backgrounds, it is assumed that not everyone could have known each other. That said, all participants actively participated throughout, particularly when it came to the discussion questions at the end of the workshop. This was also reflected during the writing component when many students chose to share their responses aloud. Writing is vulnerable in its very nature, and even more so when shared with others. This is even more true when writing about a personal topic as you implicitly add personal discovery and risk-taking into your work (Hurney 2019). Participants’ willingness to share their writing with others, even though it was not required and not noted ahead of time, demonstrated that they felt comfortable and safe enough to share vulnerable parts of themselves with the group. One of the participants commented in their survey, “I highly enjoyed the aloud reading of our reflections. I feel like it gave us a time to be truly vulnerable.” Success within this small group format (which is often a soft requirement for reflective writing groups) suggests that if implemented in an additional set of small groups, medical students could create more opportunities for community among peers and potentially have a better tolerance for vulnerability in their future practice.

One of the themes from the discussion following the reflective writing portion of the workshop was on the importance of community and the need for safe spaces where vulnerability is cultivated. Multiple participants reflected on the isolation and exhaustion that can accompany the first two years of medical school. Previous research has identified the existence of loneliness in medical professionals, including students, residents, and fellows (Keiner et al. 2023). Creating safe spaces like this workshop where trainees can reflect on vulnerable emotions and experiences and receive real time reactions/responses is therefore a potential intervention in not only the prevalence of loneliness, but also potentially burnout. The medical humanities have been previously researched as an intervention to reduce burnout and improve overall mental health within healthcare (Mangione et al. 2018, Narayan et al. 2018, Wald et al. 2019). These ideas were echoed in this workshop where several participants noted how nice it was to have this as a break from their normal class schedule. During the discussion, individuals noted the importance of having a space where they slow down and think about these topics. It would then potentially be beneficial to expand this work longitudinally to explore the idea of whether repetitive reflective writing practice in medical training could help reduce prevalence of burnout and loneliness in trainees.

Physicians spend a large amount of their time working alongside patients. Because of this, they must be skilled in communication, both with the various members of a patient’s care as well as with the patient themselves. An important aspect of care is helping patients to choose treatments that best align with their personal values, practices, and attitudes, it is essential for physicians to have experience with identifying values. Reflective practice, specifically reflective writing, can be helpful as a way to make space for identifying personal values, especially those that may be more implicit than explicit. Many people use writing to think and as a form of non traditional “data analysis” (Richardson and St. Pierre 2018) because of the way it can reveal new things about oneself (Henriksen et al. 2022). By encouraging future physicians to participate in exercises such as this one, we are teaching them to think in new and unique ways. Though this may introduce discomfort for some learners, it is important to place learners in safe uncomfortable situations if it means they will be better prepared to provide future care.

This workshop was limited to first and second year medical students, and was restricted to a small group of participants. Next steps would thus be to expand the audience. This would first mean including a larger number of preclinical students. It is not lost on the authors that the participatory nature of the workshop led to participant bias of its value. By expanding the workshop to include a larger audience of first and second year medical students, the study would be more generalizable to the larger medical learning community. Furthermore, we would like to explore expanding it beyond preclinical learners. Multiple faculty and more advanced students have also expressed interest in this work. This would create a new perception of the workshop as these individuals would have a much different perspective on values than early trainees. In the same regard, it would be interesting to explore a diversified session including participants of varying stages of training.

One unique aspect of this study in particular is the precedent it sets for humanities expansion within a medical learning community. Previous literature has suggested that medical students are not interested or engaged in the humanities (Shapiro 2009). The results of this workshop suggest otherwise. As a result of this work, we are expanding student-led humanities work within the College of Medicine. Though this takes many forms, three distinct avenues are a humanities journal, a community podcast, and an art feature. For the journal, current medical students are working to form a faculty review board and student advisory board in preparation for releasing our first call for submissions. For the community podcast, medical students are drafting interview questions to be used with a diverse set of faculty and administrators to help other medical students see their leadership as more than a name on an office. And for the art feature, the college’s humanities organization has received approval to create a new student-designed exhibit which will be displayed in the student lounge. These are all initiatives that would not be possible without displayed student investment in the humanities. This workshop is thus one example of concrete student interest.

## Data Availability

All data produced in the present study are available upon reasonable request to the authors

## Acknowledgments

The authors thank Isaac Krebs for his help with the literature review, and the participants in the workshop for their time and vulnerability.

## Ethical Approval

The Institutional Review Board at the University of Kentucky reviewed the study and approved it as minimal risk and an exempt study (IRB #:90813).

